# Health expenditure among the outpatient of type-2 diabetes in selected hospital of Kathmandu district: A cross sectional study

**DOI:** 10.1101/2021.05.27.21257843

**Authors:** Rasmita Shrestha, Aditya Shakya

## Abstract

**Introduction:** Out of Pocket (OOP) expenditure is the dominant financing mechanism in the low and middle-income countries. In these countries the prevalence of diabetes has been rising more rapidly which can lead to various micro-vascular complications thus increasing the risk of dying prematurely.

**Methods:** A cross-sectional - comparative and hospital-based study was carried out in which OOP expenditure of diabetic patient treating in public and private hospital was compared. A total of 154 diabetic patients i.e.77 in each type of hospitals were selected purposively in consultation with attending physician and staffs. Face to face interview was done to diabetic patient with a minimum of one year of illness using structured questionnaire. Lorentz curve and concentration curve were prepared using income and expenditure of the patients.

**Result:** Among154 patients, 97.4% patients had paid out of pocket for the treatment of diabetes. Mean direct cost per month was NRs. 7312.17 in public and NRs. 10125.31 in private hospital. Direct medical cost had higher share in total direct cost i.e. 60.5% in public and 69.3 % in private hospital. Medicine cost had higher percentage share (50.9%) in public hospital and laboratory cost had higher percentage share (68%) in private hospital.

**Conclusion:** Direct medical cost was higher in private hospital as compared to public hospital. All the income groups have to pay similar amount of money for the treatment i.e. economic burden for the treatment of disease was found higher for the poor people as there was not any financial protection mechanism.

## Introduction

Diabetes is the leading cause of death worldwide with global prevalence of 8.5%(1).Low and middle income countries currently bears the highest burden of non-communicable diseases(NCDs) (2).In 2012, nearly 3/4^th^ of NCDs deaths occurred in low- and middle-income countries with about 48% of deaths occurring before the age of 70and diabetes alone caused 1.5 million deaths(3).People with diabetes have increased risk of developing long term health complications like cardiovascular disease, blindness, kidney failure and lower limb amputation. This also leads to lifetime use of health resources with high spending on health care. The global health spending to treat diabetes and prevent complications was estimated to range from $673 billion to $1,197 billion in 2015(4).

The increasing burden of diabetes and its economic impact is also prominent in Nepal. According to Nepal Burden of disease 2017,diabetes contribute 1.85% of total DALY and 8^th^ edition of IDF(2017) suggest that 657,200 people are suffering from this disease in Nepal(5). Like other low and middle income countries(6) health financing in Nepal is highly dependent on out of pocket expenditure(7, 8) and do not have functional population wide insurance system throughout the country(9). Hence, the treatment and management of chronic condition of diabetes will impose a large economic burden on people with diabetes and their families in terms of higher out-of pocket (OOP) health-care payments(10) as well as economic burden to health system as a whole (8).OOP acts as a financial barrier for the poor people who are unable to pay for the health services(11).It further pushes households to unprecedented financial catastrophe and impoverishment(12).

Furthermore, health care system is delivered through public and private healthcare channels. Public health care is usually provided by the government through national healthcare systems. However the basic health service package provided by government have the limited services for NCDs; people have to rely on private or higher centers for services where they have to pay OOP(13).

In between 1995–1996 and 2010–2011 the average per capita OOP spending on Nepalese health in had increased 7 times in nominal terms. Due to the health financing system of Nepal being regressive, higher spending for healthcare are done by poorer households, as a result of which 13% of all households were found to bear the catastrophic health expenses in 2010–2011(14). Another study in Kathmandu valley showed that about 13% of household suffer from catastrophic health expenditure by paying out of the pocket for the use of healthcare(15).It provided evidence relating diabetes illnesses to catastrophic out-of-pocket expenditure on health care.

Knowledge of the healthcare cost borne by diabetics is a key ingredient for improving the quality of the management of the disease. It will also help health policymakers to map out better and more effective educational, preventive and disease management campaigns(16). However, in developing countries the information on the price that patient pay/cost from patient perspective for treating and managing diabetes is not ample(17). In the context of Nepal I have found only few published studies that have estimated the cost of illness of DM.

So, this study was carried out to compare the out of pocket expenditure among the outpatient of type-2 diabetes in selected public and private health facilities of Kathmandu district.

## Methods

### Study design and study area

It was a cross-sectional - comparative and hospital based study carried out purposively in one public and one private hospital of Kathmandu district with identical service i.e. same consulting physician in both hospitals and where mostly referred patients visit (April-Nov 2017).

### Study population

Sample size was calculated by using the method of comparing two means and it came out to be 77 for each hospital. Public hospital provides diabetic service thrice a week where as private one provides throughout the week.

Using this formula of comparison of two means,

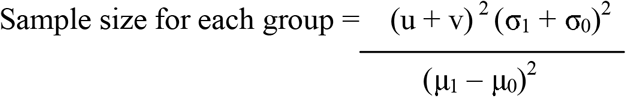

For this study, all the data are taken from the previous cost related study for type-2 diabetes held in Kathmandu valley (18). Total sample size including 8% non response was 153.36=154

### Data collection tools and technique

A pre-tested semi-structured questionnaire was used for data collection, which was adapted from a similar study held in Kathmandu valley(18)..For data collection, patients in the waiting room in both public and private hospital were approached in consultation with attending physician and staffs. Each of the eligible patients visiting during the period of study (April-Nov 2017) were interviewed until the sample size was met. Patients who were newly diagnosed with diabetes (less than a year of illness) and women with gestational diabetes during the study period were not included.

The direct expense (direct medical and direct non-medical cost) of diabetic patients over the 30 days before the study period was reported through face to face interview. The expenditure calculated in this study is based on the self-reporting by the patients. The laboratory tests cost and doctors fee was verified by checking previous bills wherever possible. Data collection was entirely done by researcher using the technique of face to face interview.

Socio-demographic factors, income of the family, type of health facility visited, number of years with disease etc. were also assessed by using questionnaire.

### Outcome assessed

Direct Medical Cost: The expenditure on consultation with doctor, medicine like insulin, lab investigations were included in direct medical cost.

Direct Non-Medical Cost: The expenditure on travel to reach the health facility, food and accommodation were included in direct non-medical cost.

### Data Analysis

Data entry was done by using Epi-data and analysis was carried out using IBM SPSS software version 21. Since, the total direct cost was not normally distributed among the respondents; non-parametric tests (Mann-Whitney U test and Kruskal-Wallis test) were used.

In this study, Lorentz curve was prepared by plotting ranks of the respondent in the x-axis and cumulative sum of Income (I) / Total income (I) of the respondents in y-axis. Concentration curve was obtained by plotting income ranks of the respondent in the x-axis and Direct Cost/Total Direct Cost in y-axis.

### Ethical consideration

Ethical clearance for the study was obtained from the Institutional Review Board of Maharajgunj Medical Campus. Verbal consent was taken from the director and doctors of both hospitals prior to data collection. Verbal and written informed consent was taken from the respondents prior to interview.

## Results

### Socio-demographic variables

The mean age of respondent was 54.56 years. Majority (55.2%) of the respondents were female. About 57.1% patients were the usual resident of Kathmandu valley while 28.6% were from urban area outside the valley and 14.3 were from rural area outside the valley. Of all the patients, 92.9%were married. Among the patients interviewed, majority (35.7%) were Janajati. In public hospital most of the patients were illiterate (32.5%) while in private 28.6% had done bachelor.

**Table 1:**
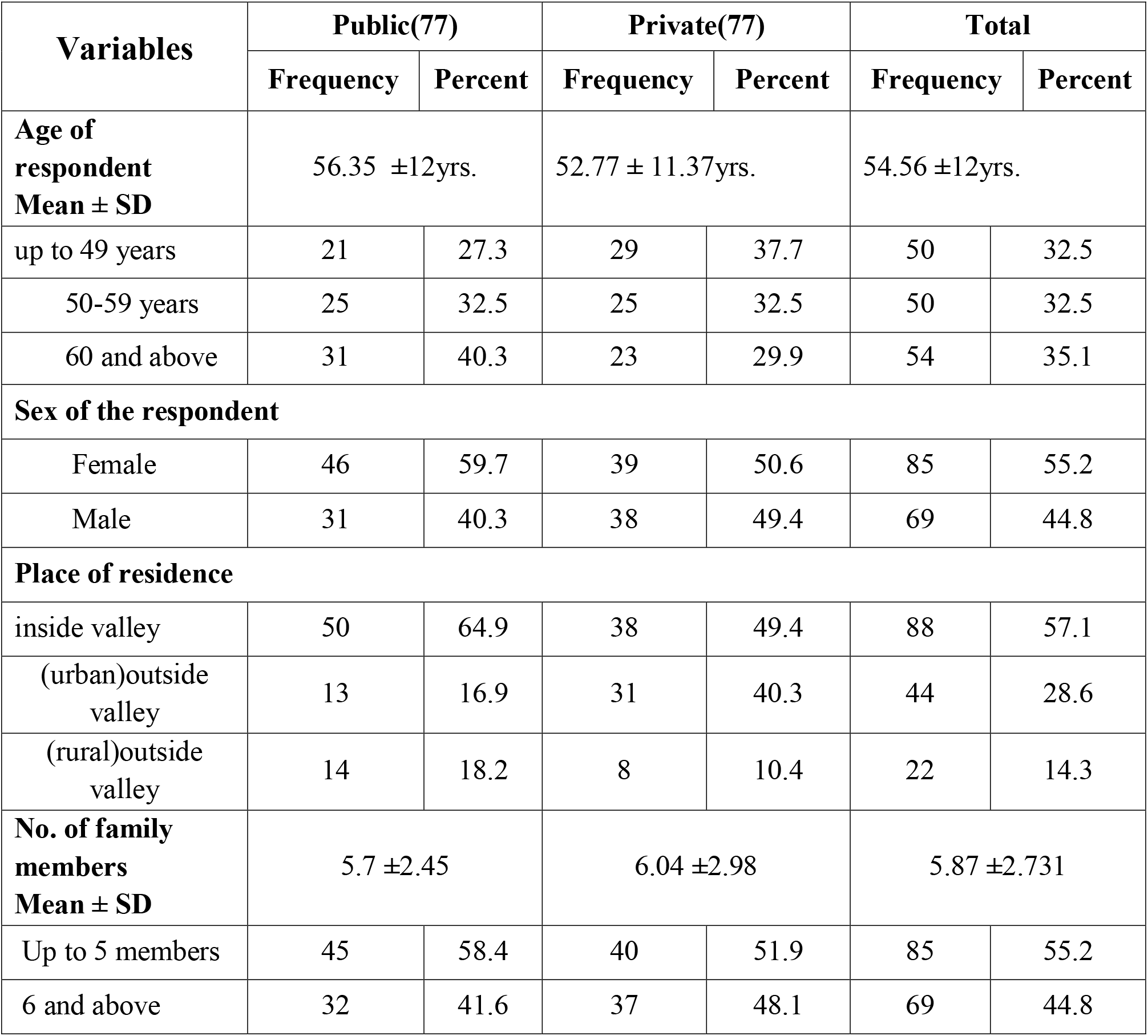
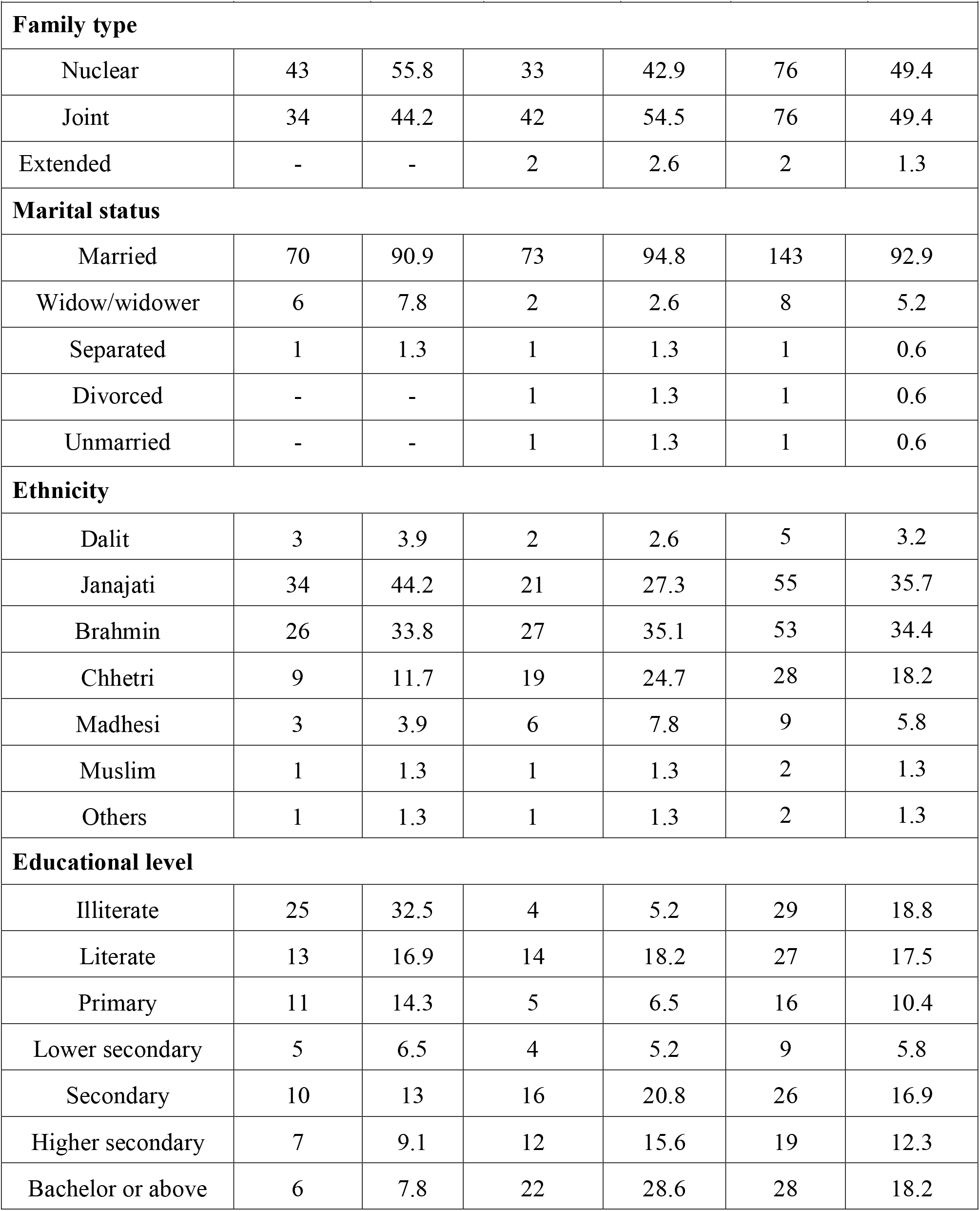
Socio-demographic variables.

| **Variables** | **Public(77)** |  | **Private(77)** |  | **Total** |  |
| --- | --- | --- | --- | --- | --- | --- |
|  | **Frequency** | **Percent** | **Frequency** | **Percent** | **Frequency** | **Percent** |
| **Age of respondent**<br>**Mean ± SD** | 56.35 ±12yrs. | | 52.77 ± 11.37yrs. | | 54.56 ±12yrs. | |
| up to 49 years | 21 | 27.3 | 29 | 37.7 | 50 | 32.5 |
| 50-59 years | 25 | 32.5 | 25 | 32.5 | 50 | 32.5 |
| 60 and above | 31 | 40.3 | 23 | 29.9 | 54 | 35.1 |
| **Sex of the respondent** |  |  |  |  |  |  |
| Female | 46 | 59.7 | 39 | 50.6 | 85 | 55.2 |
| Male | 31 | 40.3 | 38 | 49.4 | 69 | 44.8 |
| **Place of residence** |  |  |  |  |  |  |
| inside valley | 50 | 64.9 | 38 | 49.4 | 88 | 57.1 |
| (urban)outside valley | 13 | 16.9 | 31 | 40.3 | 44 | 28.6 |
| (rural)outside valley | 14 | 18.2 | 8 | 10.4 | 22 | 14.3 |
| **No. of family members**<br>**Mean ± SD** | 5.7 ±2.45 | | 6.04 ±2.98 | | 5.87 ±2.731 | |
| Up to 5 members | 45 | 58.4 | 40 | 51.9 | 85 | 55.2 |
| 6 and above | 32 | 41.6 | 37 | 48.1 | 69 | 44.8 |

| Variables | Public(77) |  | Private(77) |  | Total |  |
| --- | --- | --- | --- | --- | --- | --- |
|  | Frequency | Percent | Frequency | Percent | Frequency | Percent |
| **Family type** |  |  |  |  |  |  |
| Nuclear | 43 | 55.8 | 33 | 42.9 | 76 | 49.4 |
| Joint | 34 | 44.2 | 42 | 54.5 | 76 | 49.4 |
| Extended | - | - | 2 | 2.6 | 2 | 1.3 |
| **Marital status** |  |  |  |  |  |  |
| Married | 70 | 90.9 | 73 | 94.8 | 143 | 92.9 |
| Widow/widower | 6 | 7.8 | 2 | 2.6 | 8 | 5.2 |
| Separated | 1 | 1.3 | 1 | 1.3 | 1 | 0.6 |
| Divorced | - | - | 1 | 1.3 | 1 | 0.6 |
| Unmarried | - | - | 1 | 1.3 | 1 | 0.6 |
| **Ethnicity** |  |  |  |  |  |  |
| Dalit | 3 | 3.9 | 2 | 2.6 | 5 | 3.2 |
| Janajati | 34 | 44.2 | 21 | 27.3 | 55 | 35.7 |
| Brahmin | 26 | 33.8 | 27 | 35.1 | 53 | 34.4 |
| Chhetri | 9 | 11.7 | 19 | 24.7 | 28 | 18.2 |
| Madhesi | 3 | 3.9 | 6 | 7.8 | 9 | 5.8 |
| Muslim | 1 | 1.3 | 1 | 1.3 | 2 | 1.3 |
| Others | 1 | 1.3 | 1 | 1.3 | 2 | 1.3 |
| **Educational level** |  |  |  |  |  |  |
| Illiterate | 25 | 32.5 | 4 | 5.2 | 29 | 18.8 |
| Literate | 13 | 16.9 | 14 | 18.2 | 27 | 17.5 |
| Primary | 11 | 14.3 | 5 | 6.5 | 16 | 10.4 |
| Lower secondary | 5 | 6.5 | 4 | 5.2 | 9 | 5.8 |
| Secondary | 10 | 13 | 16 | 20.8 | 26 | 16.9 |
| Higher secondary | 7 | 9.1 | 12 | 15.6 | 19 | 12.3 |
| Bachelor or above | 6 | 7.8 | 22 | 28.6 | 28 | 18.2 |

### Socio-economic variables

Regarding family’s main source of income, most (23.4%) were involved in private job. In public hospital majority of the patients i.e. 67.5% had family’s income within a range of NRs. 10,001-50,000 and in private hospital 57.1% had income above NRs.50, 000. Also the mean monthly income of the patient’s family visiting to the public hospital (NRs. 39590.91) was lower than that of those visiting to the private hospital (NRs.82, 390.91).

**Table-2:** socio-economic variables.

| Variables | Public(77) |  | Private(77) |  | Total |  |
| --- | --- | --- | --- | --- | --- | --- |
|  | Frequency | Percent | Frequency | Percent | Frequency | Percent |
| **Family's main source of income** |  |  |  |  |  |  |
| Agriculture | 16 | 20.8 | 5 | 6.5 | 21 | 13.6 |
| Labor | 7 | 9.1 | 1 | 1.3 | 8 | 5.2 |
| Business | 10 | 13 | 24 | 31.2 | 34 | 22.1 |
| Government service | 9 | 11.7 | 23 | 29.9 | 32 | 20.8 |
| Private | 17 | 22.1 | 19 | 24.7 | 36 | 23.4 |
| Foreign employment | 14 | 18.2 | 5 | 6.5 | 19 | 12.3 |
| Others | 4 | 5.2 | - | - | 4 | 2.6 |
| **Monthly income**<br>**Mean ± SD** | Rs.39590.91±29935.75 | | Rs.82390.91 ± 81236.39 | | Rs.60990.91 ± 6468.55 | |
| less than 10000 | 12 | 15.6 | 1 | 1.3 | 13 | 8.4 |
| 10001-50000 | 52 | 67.5 | 32 | 41.6 | 84 | 54.5 |
| above 50000 | 13 | 16.9 | 44 | 57.1 | 57 | 37 |

### Diabetes related variables

Majority of the patients belong to the group less than or equal to 10 years of illness i.e. 66.2% in public and72.7% in private hospital. In public hospital about 51% of the patient had co-morbidity where as in private it was 40.3%. Among them, 82.9% had hypertension followed by thyroid (17.1%) and hypotension (5.7%). About 19% patients had complications due to diabetes. In public majority of the patients had cardiovascular disease (38.9%) and in private most of the patients had foot complication (54.5%).In public hospital 42.9% patients had paid for sugar free products like sugar-free tablets, wheat flour, boil rice etc. while in private it was 54.5%.

**Table-3:** Diabetes related variables.

| Variables | Public(77) |  | Private(77) |  | Total |  |
| --- | --- | --- | --- | --- | --- | --- |
|  | Frequency | Percent | Frequency | Percent | Frequency | Percent |
| **No. of years with diabetes** | Median=5<br>IQR=2-12yrs. |  | Median=7<br>IQR=2-12yrs. |  | Median=6<br>IQR=2-12yrs. |  |
| ≤10 years | 51 | 66.2 | 56 | 72.7 | 107 | 69.5 |
| >10 years | 26 | 33.8 | 21 | 27.3 | 47 | 30.5 |
| **Co-morbidity (multiple)** | 39 | 50.6 | 31 | 40.3 | 70 | 45.5 |
| Hypertension | 36 | 92.3 | 22 | 71.0 | 58 | 82.9 |
| Hypotension | 1 | 2.6 | 3 | 9.7 | 4 | 5.7 |
| Thyroid | 4 | 10.3 | 8 | 25.8 | 12 | 17.1 |
| **Complications (multiple)** | 18 | 23.4 | 11 | 14.3 | 29 | 18.8 |
| Cardiovascular | 7 | 38.9 | 1 | 9.1 | 8 | 27.6 |
| Retinopathy | 5 | 27.8 | 3 | 27.3 | 8 | 27.6 |
| Depression | 2 | 11.1 | 0 | 0 | 2 | 6.9 |
| Nephropathy | 2 | 11.1 | 2 | 18.2 | 4 | 18.8 |
| Neuropathy | 3 | 16.7 | 1 | 9.1 | 4 | 13.8 |
| Foot complications | 4 | 22.2 | 6 | 54.5 | 10 | 34.5 |
| **Paid for sugar free products** | 33 | 42.9 | 42 | 54.5 | 75 | 48.7 |

IQR= Inter-quartile range

### Cost related variables

In public health facility 94.8% patients had paid out of pocket for the treatment of diabetes while remaining patients were supported by government and INGOs for the payment. In private health facility all of the patients paid out of pocket for the treatment.

**Table-4:** OOP expenditure for treatment

| Variable | Public(77) |  | Private(77) |  | Total |  |
| --- | --- | --- | --- | --- | --- | --- |
|  | Frequency | Percent | Frequency | Percent | Frequency | Percent |
| paid out of pocket for the treatment | 73 | 94.8 | 77 | 100 | 150 | 97.4 |

### Total direct cost

On an average a patient spent NRs. 7312.17 for the monthly treatment in public hospital whereas the figure for private hospital was NRs. 10125.31. Diabetic patient had to pay 16.1% higher direct costs for treatment in private hospital in the last 30 days.

**Table 5:** monthly total direct cost spent in public and private hospitals

|  | N | Minimum | Maximum | Mean | Std. Deviation |
| --- | --- | --- | --- | --- | --- |
| Public | 77 | 790 | 91110 | 7312.17 | 11263.489 |
| Private | 77 | 925 | 64425 | 10125.31 | 10210.193 |

All the cost were calculated in Nepalese rupees, for standardization 1US\$ = NRs. 104.15 can be considered

### Overview of direct medical and non-medical costs

Direct medical costs had higher share in total direct cost than direct non-medical cost i.e. 60.5% in public and 69.34 % in private hospital. Mean direct medical cost was more than two times higher in private hospital as compared to public.

**Table-6:** Overview of direct medical and non-medical costs in public and private hospitals.

| Health facility | Component | N | Mean (in NRs) | Median (in NRs) | Q1 | Q3 | SD | % Share Total cost | 95% CI |
| --- | --- | --- | --- | --- | --- | --- | --- | --- | --- |
| Public | Direct medical cost | 77 | 2820.2 | 1587 | 997.5 | 3730 | 2715.4 | 60.5 | 2203.9-3436.6 |
|  | Direct non-medical cost | 77 | 1840.6 | 60 | 50 | 1600 | 4044.3 | 39.5 | 922.7-2758.6 |
| Private | Direct medical cost | 77 | 6234.8 | 5525 | 4035 | 7285 | 3708.1 | 69.4 | 5393.1-7076.4 |
|  | Direct non-medical cost | 77 | 2753.2 | 400 | 60 | 1650 | 6591.9 | 30.6 | 1257-4249.4 |

**Table-7:** Cost components of direct medical cost. Of all the components of direct medical cost, medicine cost had higher percentage share (50.9%) in public hospital and laboratory cost had higher percentage share (68%) in private hospital.

| Components of Direct medical cost | Public(77) |  | Private(77) |  | Total |  |
| --- | --- | --- | --- | --- | --- | --- |
|  | Mean (in NRs) | % share of direct medical cost | Mean (in NRs) | % share of direct medical cost | Mean (in NRs) | % share of total direct medical cost |
| Consultation cost | 37.8 | 1.3 | 568.8 | 9.1 | 303.3 | 6.6 |
| Laboratory cost | 1375.7 | 47.8 | 4242.2 | 68 | 2808.9 | 61.6 |
| Medicine cost | 1463.8 | 50.9 | 1423.7 | 22.8 | 1443.3 | 31.6 |

**Table-8:** Cost component of direct non-medical cost. Among all the components of direct non-medical cost, food and accommodation cost had higher percentage share in both public and private hospital i.e. 78.7% and 76.5% respectively. Accommodation for the people living in Kathmandu valley was considered zero for this study.

| Components of Direct non-medical cost | Public(77) |  | Private(77) |  | Total |  |
| --- | --- | --- | --- | --- | --- | --- |
|  | Mean (in NRs.) | % share of direct non-medical cost | Mean (in NRs.) | % share of direct non-medical cost | Mean (in NRs.) | % share of total direct non-medical cost |
| Travel cost | 1484.8 | 21.3 | 2354.5 | 23.5 | 1919.7 | 22.9 |
| *Food and accommodation cost | 5480 | 78.68 | 7675 | 76.52 | 6455.6 | 77.1 |

\*n=5 for public and n=4 for private
All the travel cost are measured in terms of cost to be paid for transportation in public vehicle

### Association tests

#### Association of direct cost with no. of years with diabetes, sex of the respondent and type of health facility

The Mann-Whitney U test showed that there is significant difference in the total direct costs to be paid by the patients with number of years of diabetes meaning that as the number of year with disease increases the amount of total direct cost to be paid also increases. It was found that total direct cost was significant with sex of the respondent and type of health facility. The mean direct cost was higher for males compared to females i.e. 8800 vs. 8652.8and diabetic patient had to pay higher direct costs for treatment in private hospital in the last 30 days.

**Table-9:** Association of direct cost with no. of years with diabetes, sex of respondent and type of health facility.

| Variables | N | Mean Rank | Rank Sum | z-value | p-value |
| --- | --- | --- | --- | --- | --- |
| No. of years with diabetes |  |  |  |  |  |
| ≤10 years | 107 | 72.54 | 7762 | -2.081 | 0.037* |
| >10 years | 47 | 88.79 | 4173 |  |  |
| Sex of the respondent |  |  |  |  |  |
| Female | 85 | 69.74 | 5927.5 | -2.398 | 0.016* |
| Male | 69 | 87.07 | 6007.5 |  |  |
| Health facility type |  |  |  |  |  |
| Public | 77 | 62.95 | 4847.5 | -4.047 | 0.00* |
| Private | 77 | 92.05 | 7087.5 |  |  |

#### Association of direct cost with monthly income of the patient’s family

The kruskall-wallis test showed that total cost was significantly different according to the place of residence. However, there was no any significant difference in the distribution of total direct cost and family’s monthly income. So, the entire income group paid similar amount of money for the treatment.

**Table 10:** Association of direct cost with place of residence and monthly income of the patient’s family.

| Variables | N | Mean Rank | Chi –square (df) | P-value |
| --- | --- | --- | --- | --- |
| Place of residence |  |  |  |  |
| inside valley | 88 | 63.78 | 20.016(2) | 0.001* |
| (urban)outside valley | 44 | 98.78 |  |  |
| (rural)outside valley | 22 | 89.80 |  |  |
| Monthly income |  |  |  |  |
| less than 10000 | 13 | 72.19 | 3.765(2) | 0.152 |
| 10001-50000 | 84 | 72.15 |  |  |
| above 50000 | 57 | 86.60 |  |  |

#### Distribution of burden of Direct Medical Cost across income groups

Lorentz curve was below the line of equality indicating high income inequality. Nearly 45% of the low income population (x axis) holds 20% of total respondent’s income (y axis). While remaining 55 percent of the high income group holds 80% of the respondent’s income. However, relative burden of direct medical cost (ratio of cost to income) was more among the lower income group. About 45% of lower income group (which holds only 20% of respondent’s income) had beard nearly 45% of total direct medical cost (concentration curve) in private hospital where as in public hospital it was about 40%. This indicates that out of pocket expenditure was not corresponded to the income distribution of the respondents also cost burden was slightly lower in public hospital as compared to the private ones.(fig-1 and fig-2)

**Figure 1:**
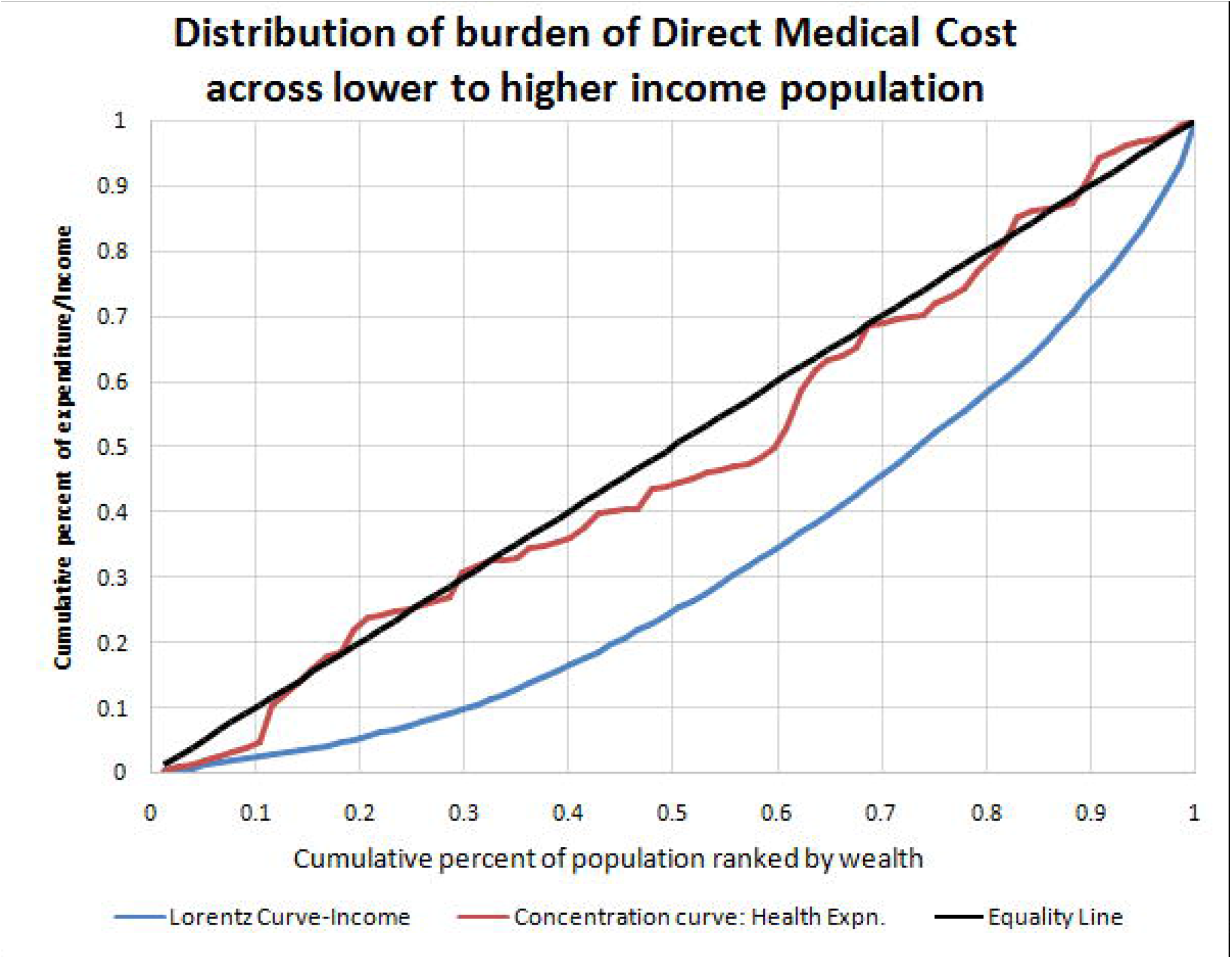
Distribution of burden of Direct Medical Cost across lower to higher income population in public hospital.

**Figure 2:**
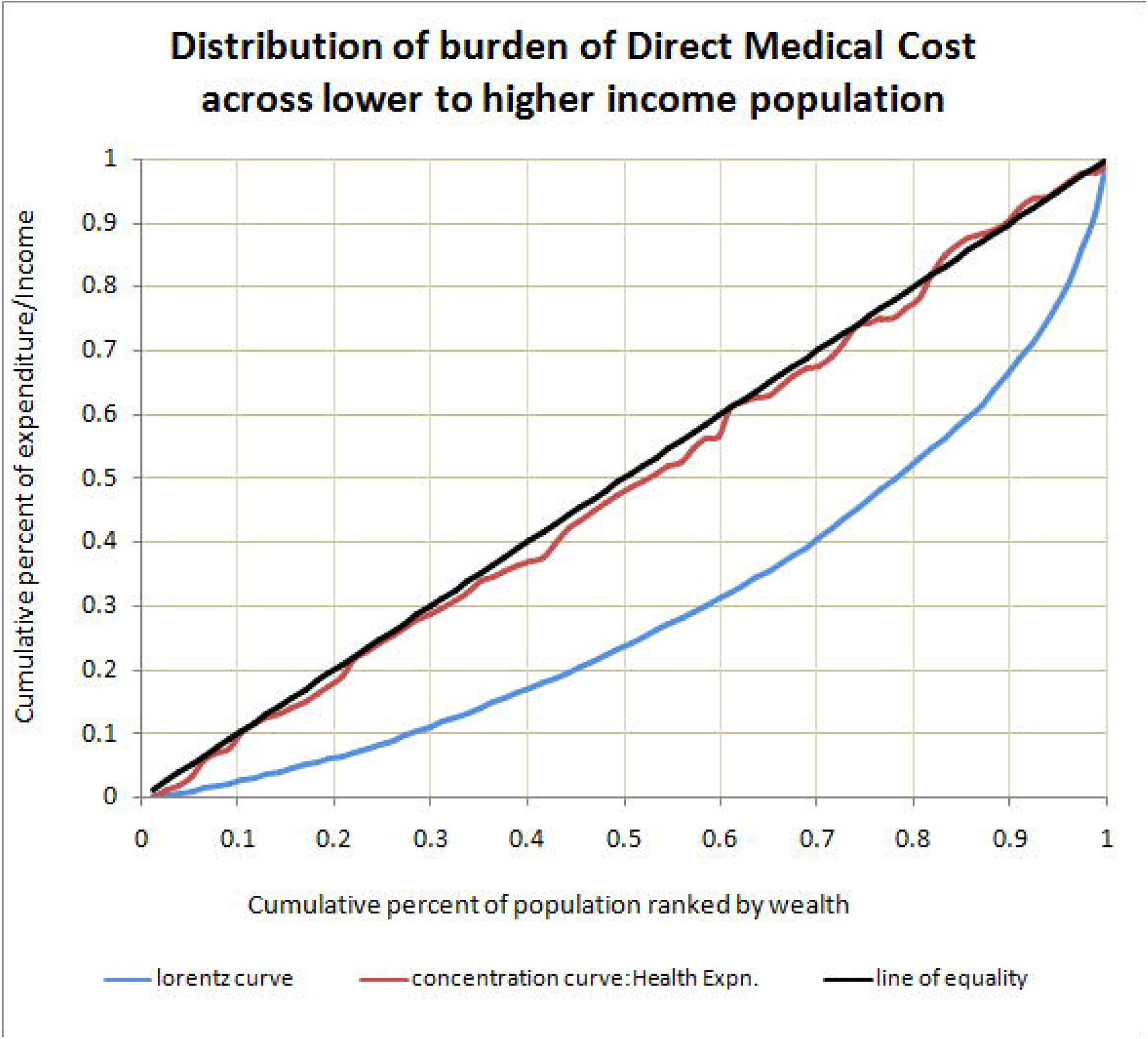
Distribution of burden of Direct Medical Cost across lower to higher income population in private hospital.

## Discussion

This study compared the out of pocket expenditure of the diabetic outpatients in public and private hospitals of Kathmandu district. Firstly, it indicates how much society is spending on diabetes care, which can then be weighed against the cost of implementing prevention programs. Secondly, it identified the different components of direct cost and the magnitude of the contribution of each component. Thirdly, this study has recognized the cost of diabetes care in relation to different socio-demographic and socio-economic characteristics.

This study indicates that 97.4% people had to pay out of pocket for the treatment remaining respondents were supported by government and various INGOs for the treatment as some of them were government employee and some were refugee. As per Grover and colleagues similar percentage of cost is paid by patients and their families in India(19).This suggest that social protection does not exist virtually as most expenses are out-of-pocket (20). The economic impact of high out of pocket payment for management of diabetes at household level(21) and its consecutive effect on diminishing standard of life has been well established. So, there is a strong need to develop different health insurance schemes mainly targeting for the poorer segment of the population in order to protect their household budget and increase treatment compliance, which will help prevent unnecessary complication(s).

On average a person with diabetes spends NRs. 7312.17for monthly treatment in public hospital whereas this figure was NRs. 10125.31 for those visiting private hospital. This cost is substantially high for people in Nepal where 21.6% of population still lie under the poverty line (22). Furthermore, diabetic patient had to pay higher direct costs in private hospital as compared to public hospital similar was finding in one of the study conducted by Niraj and colleagues in Nepal(18). This is mainly because in Nepal, private hospital are run mainly for profit and public health care is usually provided by the government through national healthcare systems where the cost are subsidized by the government and have the provision of free basic health service(23). The major contributor for the total direct cost was direct medical cost meaning that people have to pay more for doctor fee, laboratory test and medicine than direct non-medical cost like transport and food and accommodation cost which conclude the result found in the study conducted in Pakistan and Brazil (24, 25). A study has shown that the cost of insulin for management of diabetes is highly unaffordable for majority of its population in Nepal (26).Patients who visited private hospital had to pay nearly more than thrice laboratory cost as compared to its counterparts in public hospital. Reasons for this huge difference in the cost were higher price in private hospital for the similar test and also patients were subjected to do more number of laboratory tests in private hospital. Analogous result was seen in one of the study conducted in India saying that private laboratories typically increase cost 4–7-fold than the actual reagent price(27).

It was found that patients attending the private hospital were economically better off than those visiting the public hospital with higher mean monthly family income i.e. NRs.82390.91 of those visiting private hospital and NRs. 39590.91 of those visiting public hospital. Public health services are considered to be of low quality, lower income groups use them more than higher income groups. However, the higher income groups choose private sector care as it has shorter waiting periods, longer or more flexible opening hours, and better availability of staff(28, 29).Another finding was that, the total direct cost of diabetes care was significantly higher in patients with longer duration (≤10years vs. >10 years). This pattern was also observed in other studies conducted in Southern India and in Sweden (30, 31).As diabetes is associated with many complications, longer duration of the disease makes the condition worse which eventually results in higher treatment cost.

The percentage of patients with co-morbidity was higher in public hospital than that of private. One of the reasons for this may be the public hospital selected for the study was tertiary level hospital where most of the patients with serious health problems were referred for the treatment.

One of the interesting finding of the study was that no any significant difference exist between the total cost incurred by the patient and the family’s monthly income i.e. all the income group paid similar amount of payment for the treatment. So, the relative burden of direct medical cost (ratio of cost to income) was found more among the lower income group meaning that the same disease may not be of greater economic impact for rich people but it may have catastrophic and impoverishing impact for the poor. Similar pattern of expenditure was shown in the studies conducted in India (30, 32)

One of the strengths of the study is the comparative cost related study which provides information about the variation in OOP payment that diabetic outpatients have to pay in public and private hospital. Another one is, it can help to draw the attention of policy maker for making the health policy in regards of poor people by the effective implementation of risk pooling mechanism.

There are various limitations that need to be considered. The cost of co-morbidities and indirect cost were not included meaning that the costs are likely to be underestimated. This study is based on self-reported spending (only few have bills of their expenditure) so, there may be over or under estimation of health care payment. Due to the smaller sample size representing small geographical area and purposive nature of sampling the cost estimation may not be generalizable. Despite its limitation the study has been able to highlight some key findings related to the expenditure in diabetes that should be taken in consideration for systematic healthcare planning for diabetes care.

## Conclusion

This study suggest that OOP expenditure for treating diabetes is higher in Nepal especially for those visiting the private hospital and mostly affect the low income level people. So, some form of financial protection scheme is needed to protect them from financial risk. Similarly, diabetes is a complex disease the key to reducing costs seems to be intensive management and control in order to prevent and delay the associated late complications. Knowing its complexity of nature the government should develop a comprehensive action plan to tackle diabetes and other NCDs. In Nepal there is limited information regarding cost estimation of disease thus, more emphasis should be given for such studies in the coming days.

## Data Availability

Yes - all of the data of this research are fully available without restriction

